# Selection of AVP-shortage patients as candidates for low-dose oral desmopressin administration

**DOI:** 10.1101/2020.09.12.20188763

**Authors:** Takumi Takeuchi, Kazuki Maki, Yumiko Okuno, Mami Hattori-Kato, Koji Mikami

**Affiliations:** Department of Urology, Japan Organization of Occupational Health and Safety, Kanto Rosai Hospital; Department of Urology, Tokyo Teishin Hospital, 2-14-23, Fujimi, Chiyoda-ku, Tokyo 102-8798, Japan

**Keywords:** AVP, osmolality, desmopressin, urination

## Abstract

**Introduction:** Diabetes insipidus (DI) is characterized by the excretion of large volumes of hypotonic urine and thirst due to an impaired ability to concentrate urine, leading to uncontrolled diuresis, which may cause life-threatening dehydration and electrolyte imbalances. Central DI is caused by the deficient secretion of the posterior pituitary antidiuretic hormone arginine vasopressin (AVP). Desmopressin (Deamino-8-D-AVP, the synthetic analogue of AVP, Minirinmelt®) is generally used to treat central DI. Desmopressin orally disintegrating tablets are recently administered to male patients with nocturia. We herein attempted to select male patients with an elevated nocturnal urinary frequency possibly due to a shortage of AVP. These patients may be good candidates for low-dose oral desmopressin administration.

**Patients and methods:** Serum and spot urine osmolality, electrolytes, serum creatinine, casual blood glucose, plasma brain natriuretic polypeptide (BNP), and plasma AVP were measured at the same time in 97 elderly male patients with urinary symptoms under free water drinking. The International Prostate Symptom Score, Overactive Bladder Symptom Score, and frequency-volume charts at least twice were also evaluated.

**Results:** A binary plot of plasma AVP and serum osmolality indicated a region at which patients had relatively lower AVP considering higher serum osmolality. It was tentatively named the Desmopressin region. Twenty out of 97 (20.6 %) patients were in the Desmopressin region.

No significant differences were observed in the frequency of administered urinary drugs or existing co-morbidities between patients in the Desmopressin and non-Desmopressin regions. Daily urine output did not exceed 3 L in any patient. Plasma AVP was lower, while serum osmolality and serum sodium were higher in patients in the Desmopressin region than in those in non-Desmopressin region. Furthermore, urine osmolality was slightly lower in patients in the Desmopressin region. No significant differences were observed in urine volume, urinary frequency, or urination questionnaire scores between both groups.

**Conclusion:** AVP-shortage patients may be selected for treatment with oral desmopressin based on measurements of serum osmolality and plasma AVP. After the exclusion of patients with marked hyperglycemia, decreased cardiac, or renal function, low-dose oral desmopressin may be administered to patients with an increased urine output, nocturia, elevated plasma osmolality, and relatively low plasma AVP.

## Introduction

Diabetes insipidus (DI) is characterized by the excretion of large volumes of hypotonic urine (>50 mL/kg/24 hours in adults) and thirst due to an impaired ability to concentrate urine, leading to uncontrolled diuresis, which may cause life-threatening dehydration and electrolyte imbalances (1,2). There are three types of DI. Central DI is caused by the deficient secretion of the posterior pituitary antidiuretic hormone arginine vasopressin (AVP). The intracranial causes of central DI are neoplastic, vascular, trauma, inflammatory, infection, and idiopathic (1,2). Nephrogenic DI occurs when the response of renal tubules to absorb water is impaired due to resistance to the effects of AVP on its receptors (1,2). Gestational DI is transient during pregnancy due to an increase in the metabolism of vasopressin induced by placental cysteine aminopeptidase (1,3). Primary polydipsia is a disorder caused by excessive water intake and is often observed in patients with psychogenic issues. DI and primary polydipsia are collectively referred to as polyuria-polydipsia syndromes.

The AVP gene is in chromosome 20p13. Pre-pro-vasopressin, a peptide of 164 amino acids, is transcribed from the gene in magnocellular neurons in the supraoptic and paraventricular nuclei of the hypothalamus. It is converted to pro-vasopressin by the removal of the signal peptide and N-linked glycosylation of copeptin. AVP, neurophysin II, and copeptin are produced by the further processing of pro-vasopressin followed by axonal transportation to the posterior pituitary (2).

A water deprivation test followed by the administration of desmopressin (Deamino-8-D-AVP, the synthetic analogue of AVP, Minirinmelt®) may theoretically differentiate between central and nephrogenic DI as well as primary polydipsia; however, overlapping test results sometimes affect the accuracy of the diagnosis because of the existence of partial central DI, partial nephrogenic DI, and chronic primary polydipsia (4). The measurement of plasma AVP levels in combination with the water deprivation test has also been suggested in order to distinguish between the various polyuria-polydipsia syndromes (5). DI patients need to drink a sufficient amount of fluids to avoid dangerous dehydration. Desmopressin is usually administered to treat central DI.

Desmopressin orally disintegrating tablets are administered to male patients with nocturia (6,7). We herein attempted to select male patients with an elevated nocturnal urinary frequency possibly due to a shortage of AVP. These patients may be good candidates for low-dose oral desmopressin administration.

## Patients and methods

Serum and spot urine osmolality, electrolytes, serum creatinine, casual blood glucose, plasma brain natriuretic polypeptide (BNP), and plasma AVP were measured at the same time in 97 elderly male patients with urinary symptoms under free water drinking. The International Prostate Symptom Score (IPSS), Overactive Bladder Symptom Score (OABSS), and 24-hour frequency-volume charts at least twice were also evaluated. A 24-hour frequency-volume chart states the time and volume of each void and the bedtime and waking time. The nocturnal urine volume was defined as the volume of voids between bedtime and waking time plus the first-morning void, while the first-morning void was regarded as a normal diurnal voiding episode. Data recorded from frequency-volume charts were averaged.

## Statistical analysis

Age, prostate volume, biochemical data, urine volume, urinary frequency, and urination questionnaire scores were analyzed by the unpaired *t*-test. Rates of urination drug usages and co-morbidities were assessed by the two-tailed chi-squared test.

## Ethics

The present study was conducted in accordance with the Helsinki Declaration after approval of the Ethics Committee of Kanto Rosai Hospital (G2020-3). Written informed consent was received from all participants in the study.

## Competing interests

We declare that there is no conflict of interest regarding the publication of this study.

## Funding

There is no funding for this study.

## Results

Figure 1 shows a binary plot of plasma AVP and serum osmolality. The curves on the graph conveniently distinguished between central DI and renal DI/polydipsia (8). The region below this curve in addition to serum osmolality of more than 290 mOsm/kg was tentatively named the Desmopressin region. Twenty out of 97 (20.6%) patients were in the Desmopressin region.

**Figure 1.**
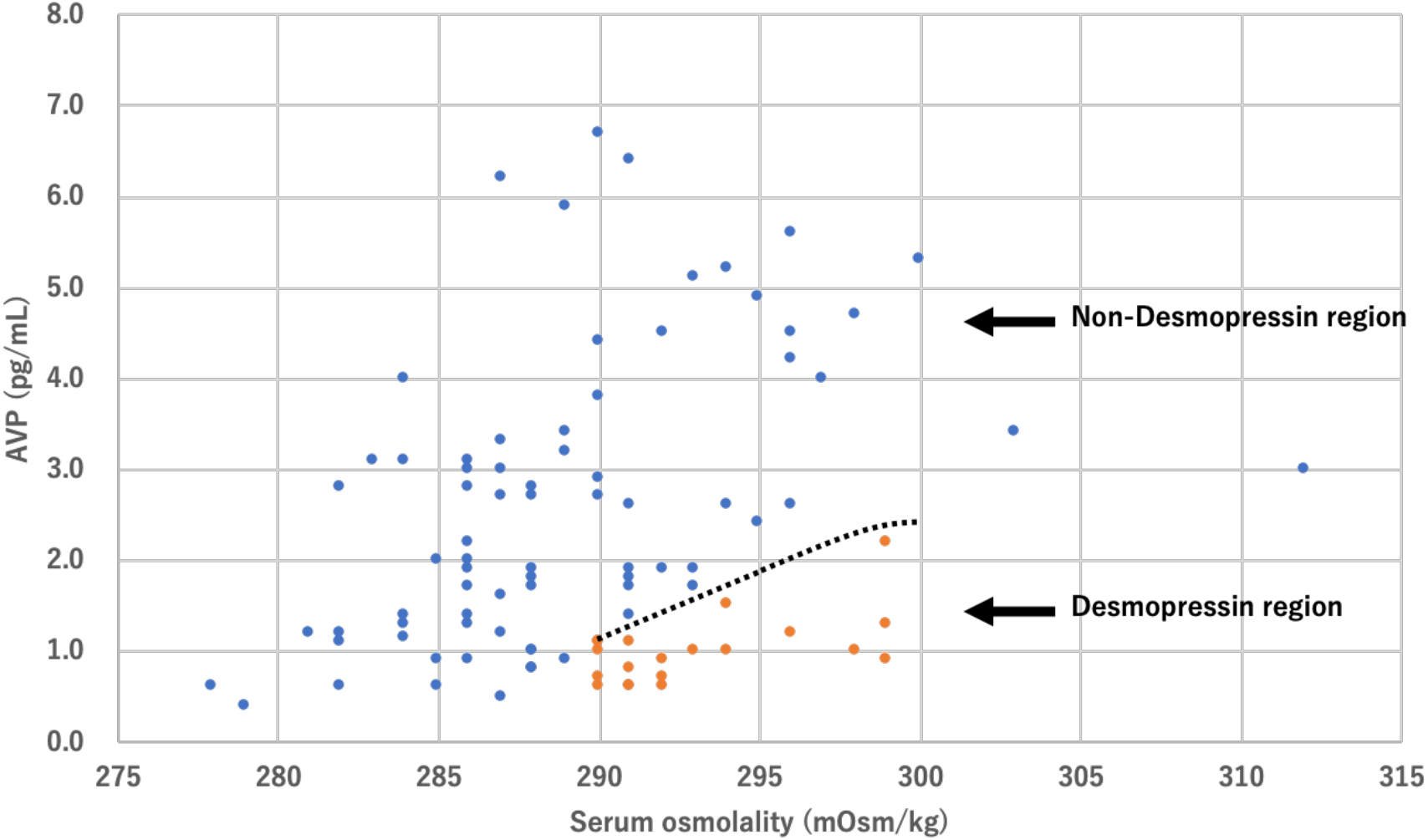
Binary plot of plasma AVP and serum osmolality in patients with urinary symptoms. Orange dots show AVP-shortage patients with elevated serum osmolality (Desmopressin region), while blue dots are non-AVP-shortage patients (non-Desmopressin region). The curve is the line that separates the two groups. Two cases showing AVP20.4 pg/mL, serum osmolality 290 mOsm/kg and AVP 10.0 pg/mL, serum osmolality 301 mOsm/kg were excluded from plotting.

Table 1 shows the urination-related drugs already administered to and co-morbidities of patients. No significant differences were observed in the frequency of administered urinary drugs and existing comorbidities in patients in the Desmopressin and non-Desmopressin regions. Table 2 shows data on biochemistry and urination. Daily urine output did not exceed 3 L in any patients. Plasma AVP was lower, while serum osmolality and serum sodium were higher in patients in the Desmopressin region than in those in the non-Desmopressin region. Furthermore, urine osmolality was slightly lower in patients in the Desmopressin region. No significant differences were observed in urine volume, urinary frequency, or urination questionnaire scores between both groups.

**Table 1:** Urinary drugs administered to and co-morbidities of patients.

|  | Non-Desmopressin region | Desmopressin region | p-value |
| --- | --- | --- | --- |
| <u>Urination drugs</u> |  |  |  |
| α1-adrenergic antagonist | 90.9 (%) | 90.0 (%) | 0.9006 |
| PDE5 inhibitor | 0.0 | 5.0 | 0.5072 |
| anti-cholinergic | 11.7 | 15.0 | 0.9843 |
| 5α-reductase inhibitor | 59.7 | 50.0 | 0.5950 |
| β3-adrenergic agonist | 5.2 | 0.0 | 0.6819 |
| <u>Co-morbidities</u> |  |  |  |
| hypertension | 42.9 (%) | 60.0 (%) | 0.2635 |
| diabetes mellitus | 16.9 | 20.0 | 0.7439 |
| intracranial | 11.7 | 10.0 | 0.8320 |
| cardiovascular | 24.7 | 25.0 | 0.9761 |
| liver | 1.3 | 5.0 | 0.8770 |
| kidney | 2.6 | 15.0 | 0.0954 |
| lung | 2.6 | 0.0 | 0.4664 |
| orthopedic | 11.7 | 15.0 | 0.9843 |
| malignancy | 18.2 | 20.0 | 0.8522 |

N=77 and N=20 for the non-Desmopressin and Desmopressin regions, respectively.

**Table 2:** Biochemistry and urination data of patients.

|  | Non-Desmopressin region | Desmopressin region | p-value |
| --- | --- | --- | --- |
| Age | 79.4±7.6 | 81.5±7.9 | 0.2791 |
| serum sodium (mEq/L) | 140.1±2.6 | 142.1±2.0 | 0.0302 |
| serum chloride (mEq/L) | 103.8±2.7 | 104.9±2.2 | 0.1011 |
| serum creatinine (mg/dL) | 1.00±0.34 | 1.08±0.29 | 0.2941 |
| casual blood glucose (mg/dL) | 122.5±35.9 | 121.8±36.0 | 0.9360 |
| plasma BNP (pg/mL) | 51.6±76.1 | 55.4±47.6 | 0.8355 |
| plasma AVP (pg/mL) | 2.94±2.68 | 1.00±0.38 | 0.0018 |
| serum osmolality (mOsm/kg) | 289.1±5.6 | 293.1±3.3 | 0.0031 |
| urine osmolality (mOsm/kg) | 527.3±151.2 | 456.1±166.4 | 0.0693 |
| prostate volume (mL) | 45.1±29.8 | 36.4±18.5 | 0.2172 |
| daytime urine volume (mL) | 903±325 | 950±343 | 0.6077 |
| nocturnal urine volume (mL) | 661±361 | 731±293 | 0.4732 |
| 24-hour urine volume (mL) | 1571±515 | 1681±430 | 0.4279 |
| nocturnal polyuria index (%) | 40.5±14.2 | 42.5±12.5 | 0.6032 |
| daytime urinary frequency | 7.5±2.1 | 7.3±1.7 | 0.6853 |
| night time urinary frequency | 2.4±1.5 | 2.8±2.0 | 0.3832 |
| 24-hour urinary frequency | 9.9±2.8 | 10.1±3.1 | 0.8416 |
| IPSS-T | 10.8±6.7 | 12.3±7.0 | 0.3860 |
| IPSS-V | 5.4±4.8 | 5.7±4.8 | 0.7819 |
| IPSS-S | 5.4±3.1 | 6.1±3.2 | 0.3818 |
| IPSS-Q7 | 2.5±1.2 | 2.6±1.4 | 0.9020 |
| IPSS-QOL | 3.2±1.5 | 3.6±1.5 | 0.2741 |
| OABSS | 5.2±3.5 | 5.7±3.3 | 0.5226 |

N=77 and N=20 for the non-Desmopressin and Desmopressin regions, respectively. IPSS-T: total IPSS score, IPSS-V: IPSS voiding score, IPSS-S: IPSS storage score. nocturnal polyuria index: nocturnal urine volume divided by 24-hour urine volume.

## Case

A male in their 90’s was admitted to our hospital with nocturia despite being administered an α-blocker, anti-cholinergic drug, and β3 antagonist at a nearby urologic clinic. His prostate volume was 16 g, residual urine was 18 mL, and he had thirst. Serum Na was 140 mEq/L, serum creatinine 0.89 mg/dL, blood glucose level 126 mg/dL, plasma BNP 45.0 pg/mL, plasma AVP 1.0 pg/mL, serum osmolality 298 mOsm/kg, and urine osmolality 542 mOsm/kg. Frequency-volume charts revealed a daily urine output of 2090 mL, night time urine output rate 50.2%, daily urination frequency 12 times, and night time urination frequency 4 times. These results indicated that the patient was located in the Desmopressin region. He had no severe cardiac dysfunction, renal dysfunction, marked hyperglycemia, or hyponatremia. Oral Desmopressin 25 μg/day was started, and his daily urine volume, nocturnal urine volume, IPSS score (total and storage), IPSS-Q7 score, and IPSS-QOL score had decreased 8 weeks after the initiation of this treatment, as shown in Figure 2.

**Figure 2.**
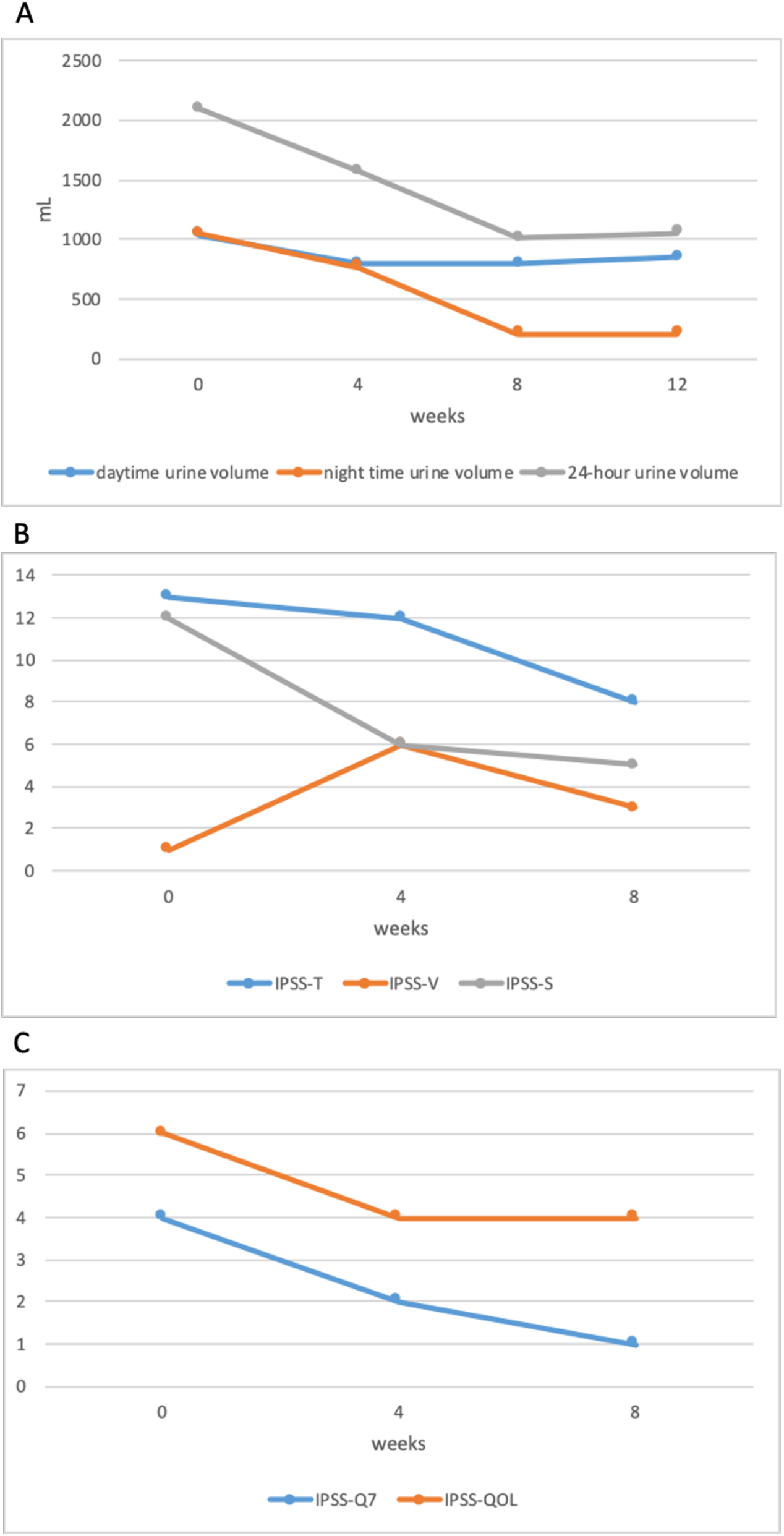
A: Changes in urine volume following the administration of Desmopressin. B: Changes in IPSS following the administration of Desmopressin. IPSS-T: total IPSS score, IPSSV: IPSS voiding score, IPSS-S: IPSS storage score. C: Changes in the IPSS-Q7 score and IPSS-QOL score following the administration of Desmopressin.

## Discussion

In the present study, none of our patients exhibited polyuria with a urine output of more than 50 mL/kg/day or 3-4 L/day, which are the diagnostic criteria for DI. However, in some patients, plasma AVP was not sufficiently elevated while plasma osmolality was higher than 290 mOsm/kg under free water drinking. Therefore, these patients appeared to have a shortage of AVP.

If AVP is low, the reabsorption of water in the renal tubules may be suppressed, causing mild dehydration and a slight increase in serum osmolality. The secretion of AVP is stimulated by an increase in serum osmolality; however, when AVP secretion from the posterior pituitary gland cannot sufficiently cope with the increased serum osmolality, the increase in plasma AVP becomes insufficient. Additionally, when urine output is increased, for example, 30 mg/kg/day or more, patients may be good candidates for low-dose oral desmopressin administration. The urinary osmolality of patients in the Desmopressin region exceeded 300 mOsm/kg in most cases, and may have been compensatory for the relative AVP shortage due to the overexpression of vasopressin V2 receptors in the renal tubules (9).

Decreases in the secretion of AVP from the posterior pituitary may be caused by intracranial diseases. In the present study, no significant differences were observed in the frequency of intracranial diseases between patients in the Desmopressin and non-Desmopressin regions, and the pathological condition directly decreasing AVP secretion was not identified. Therefore, decreased AVP secretion appeared to be idiopathic.

The development of polyuria in prominently hyperglycemic patients is due to osmotic diuresis, not a deficiency in AVP. Therefore, it is not a target of desmopressin. In addition, patients with elevated plasma BNP, indicating cardiac dysfunction and/or decreased renal function, need to be excluded because desmopressin may induce heart failure due to water overload.

Polyuria-polydipsia cases with low plasma osmolality and AVP may be caused by an increased water intake. The administration of desmopressin to these cases may induce hyponatremia and heart failure. Therefore, they are not a target for desmopressin. No significant differences were observed in urine volume, urinary frequency, or urination questionnaire scores between patients in the Desmopressin and non-Desmopressin regions. Therefore, AVP-shortage patients need to be selected for treatment with oral desmopressin based on measurements of serum osmolality and plasma AVP.

When desmopressin was administered following the selection of appropriate patients, a reduced urine volume, decreased IPSS and OABSS, improved QOL scores, and no adverse events were observed. In conclusion, after the exclusion of patients with marked hyperglycemia and decreased cardiac or renal function, low-dose oral desmopressin may be administered to patients with increased urine output, nocturia, elevated plasma osmolality, and relatively low plasma AVP.

## Data Availability

Not available

## Notes

### Competing Interest Statement

The authors have declared no competing interest.

### Clinical Trial

It is a retrospective study.

### Author Declarations

The present study was conducted in accordance with the Helsinki Declaration after approval of the Ethics Committee of Kanto Rosai Hospital (G2020-3).

